# CogGame: Gamified Cognitive Assessments in Young Adults with Suicidal Thoughts

**DOI:** 10.1101/2022.10.17.22281128

**Authors:** Christina Chae Yon Shin, Haley M LaMonica, Loren Mowszowski, Vanessa Wan Sze Cheng, Laura Kampel, Jin Han

## Abstract

**Introduction:** The susceptibility to suicidal behaviour has been linked to cognitive functioning deficits. Gamified assessments have emerged as a practical and engaging approach to assess these deficits, though their acceptability amongst young adults with suicidal thoughts is currently understudied.

**Methods:** Thirteen young Australian adults aged 18 to 25 years who experienced suicidal thoughts in the past year were recruited to evaluate the smartphone based CogGame app. Inductive thematic analysis was utilised to identify the themes obtained from the interviews. The relationships between cognitive functioning deficits and the severity of suicidal thoughts were explored by correlational analyses.

**Results:** All participants found the GogGame app easy to learn to use and navigate. Positive experiences and high user satisfaction were reported with the use of CogGame app. Major areas for improvement include having clearer instructions and app information, adjusting the difficulty of the exercises, and addressing a few technical issues such as decreasing loading time. Higher levels of suicidal thoughts were found to be significantly associated with poorer visual learning performance on the CogGame app (*p* = .01).

**Conclusion:** Positive participant experiences with CogGame revealed the promising potential of gamified assessments to measure cognitive functioning in young adults with suicidal thoughts.

## Introduction

Suicide is one of the leading causes of worldwide mortality, and has been recognised as a significant global public health issue with more efforts required to identify individuals at risk and tailor interventions to prevent suicide (World Health Organization, 2019). Well-known risk factors for suicide include a diagnosis of a psychiatric illness, a previous suicide attempt, a family history of suicide, social isolation and financial or legal difficulties (O’Connor & Nock, 2014; Turecki & Brent, 2016).

The susceptibility to suicidal behaviour has been linked to deficits in cognitive functioning (Bredemeier & Miller, 2015; Saffer & Klonsky, 2018). Evidence suggests that more severe suicidal thoughts and behaviours are associated with poorer cognitive functioning in areas including executive function (inhibition of habitual responses, switching between tasks, and incorporating new information into current thoughts) (Bredemeier & Miller, 2015; Pu, Setoyama, & Noda, 2017), memory (including short-term, long-term, autobiographical and working memory) (Keilp et al., 2014; Richard-Devantoy, Berlim, & Jollant, 2015), and attention (Ruch et al., 2020; Saffer & Klonsky, 2018). These deficits are thought to be associated with difficulty disengaging from suicidal thoughts and generating or switching to more adaptive coping strategies in response to distress (Bredemeier & Miller, 2015; Miranda, Valderrama, Tsypes, Gadol, & Gallagher, 2013), a reduced ability to envision positive outcomes (Miranda, Gallagher, Bauchner, Vaysman, & Marroquin, 2012), as well as educational and problem-solving struggles that can ultimately damage an individual’s self-perception and feelings of worth, further fuelling suicidal behaviour (Sarkisian, Hulle, & Goldsmith, 2019). Cognitive functioning deficits have thus been recognised as a potentially important modifiable risk factor for suicide that may help in identifying certain individuals, such as those without a diagnosis of depression, who may not traditionally be perceived to be at risk, as well as identifying suicidal young adults who often tend to avoid disclosing their suicidal thoughts to others (Miranda et al., 2012) and seeking help for suicidal behaviours (Aguirre Velasco, Cruz, Billings, Jimenez, & Rowe, 2020). Thus, detecting early, subtle changes in cognitive function may help identify individuals who are at risk, ultimately helping to prevent suicidal behaviour in young adults.

The emerging gamified cognitive assessments offer a potentially timely, practical, and appealing solution to detect early changes in cognitive function amongst young people. The graded challenges, appealing graphics and intuitive rules associated with games provide the inherent ability to engage the user and maintain their motivation and attention, thus improving participant experiences (Groznik & Sadikov, 2019; Lumsden, Edwards, Lawrence, Coyle, & Munafò, 2016; Potvin, Charbonneau, Juster, Purdon, & Tourjman, 2016). This is crucial in producing data with good quality and increasing the effectiveness of methods to identify individuals with poor cognitive functioning (Lumsden et al., 2016). Additionally, they may reduce test anxiety and improve ecological validity (Akoodie, 2020; Lumsden et al., 2016). However, the use and effectiveness of gamified cognitive assessments have not been adequately explored (Demant, Vinberg, Kessing, & Miskowiak, 2015; Lumsden et al., 2016; Potvin et al., 2016), and their acceptability and perceptions amongst young adults with suicidal thoughts has not been well-studied. A deeper understanding of this is imperative in potentially harnessing games as an assessment tool to identify individuals with cognitive deficits, which may be linked to a greater risk of suicide, as well as delivering possible intervention methods including cognitive training through a gamified medium (Groznik & Sadikov, 2019; Lumsden et al., 2016).

We developed a gamified cognitive assessment app (“CogGame”), incorporating the properties of digital games, to create a more engaging, accessible, and time-efficient alternative to traditional neuropsychological tests. The app, which can be accessed on a digital device such as an iPad or smartphone, was designed to promote repeated engagement, decrease test anxiety, increase ecological validity, and allow routine self-administration to track progress of cognitive functioning in conjunction with clinical management. The app consists of three assessment tasks, titled “Let’s Go Shopping (Shopping List Recall)”, “Hidden Objects (Visual Search Task)”, and “Are We There Yet? (Route Learning)”, assessing different aspects of memory and executive function within approximately 30 minutes. Though originally designed to assess cognition in older adults, we propose that this app may prove to be suitable and beneficial for younger audiences as well.

This study aims to understand how Australian young adults who experienced suicidal thoughts in the previous 12 months respond to CogGame in terms of its acceptability. A second aim was to explore the relationships between cognitive functioning assessed by CogGame and suicide ideation severity. The results from this study may further inform future research in developing effective tools for identification of suicidal ideation and behaviour in individuals, and to inform ongoing development and refinement of gamified cognitive assessments in detecting cognitive changes in younger adults.

## Methodology

### Participants

Young adults aged 18 to 25 years living in Australia, fluent in English, and who identified with having experienced suicidal thoughts in the past 12 months were recruited via emails from a research registry during September to October 2021. Participants were excluded from the study if they experienced suicidal thoughts in the seven days preceding the study, had ever attempted suicide in their lifetime, or had a diagnosis of bipolar disorder or psychosis for safety management. Among the 33 participants who completed screening assessments to confirm their eligibility, 13 participants met the eligibility criteria and completed the measures for the study.

### Data Collection Procedure

Data collection involved two-steps: a Qualtrics online survey including questions on sociodemographic and mental health related variables, followed by a video-conferencing session, during which participants completed the cognitive assessments on the CogGame app, as well as a 50-minute semi-structured interview to obtain their feedback on the acceptability of the app. Their responses were audio-recorded, transcribed verbatim by CCYS and de-identified. Informed consent was obtained from all participants prior to commencing the online questionnaires and the video-conferencing session hosted by the Microsoft^®^ Teams. Ethics approval was granted by the Human Research Ethics Committee at the University of New South Wales (HC210432), and participants were reimbursed for their time with a $60 e-gift voucher.

### Measures

#### Suicidal Thoughts

The severity of suicidal thoughts was assessed by the Suicidal Ideation Attributes Scale (SIDAS) (Van Spijker et al., 2014). The SIDAS is a five-item scale, with four items scored from 0 to 10 and one item reverse-scored, which is then totalled to produce an overall score, with a higher score indicating more severe suicidal ideation (range 0 to 50). The SIDAS demonstrated good internal consistency, with a Cronbach’s alpha of 0.90 in the current study.

#### Cognitive Performance

Quantitative data on cognitive performance was obtained using CogGame (see Figures 1 and 2), a new smartphone application developed by a group of researchers (HML, JH, LM and VWSC) from the University of Sydney and the University of New South Wales. This app consists of three gamified assessment tasks, each testing different aspects of memory and executive functioning (Table 1). For the games “Let’s Go Shopping” and “Are We There Yet”, each cognitive function assessment was scored by summing the number of correct responses selected, thus a higher score represented greater cognitive function. The second game (“Hidden Objects”) was scored by summing the number of incorrect responses selected, with a lower score representing greater visual processing speed.

**Table 1:** Outline of gamified assessments and procedure.

| Game and Procedure | Aspect of Cognitive Function Assessed | Task Instructions | Score Range |
| --- | --- | --- | --- |
| **Let's Go Shopping** |  |  |  |
| Practice round | Practice | Practice round to familiarise participants with the interface and instructions. | – |
| Rounds 1, 2, 3 | Verbal learning | In each round, participants were instructed to memorise the same 12 grocery items, presented to them one at a time as written words. They were then instructed to select the correct 12 items from a grid of 18 visually-presented items. | 0 – 36 |
| Round 4 | Verbal memory | After a short delay, participants were instructed to select the target grocery items from the same grid from memory. | 0 – 12 |
| Round 5 | Recognition memory | After further delay, participants were instructed to select the correct items from an array of incorrect and correct items from memory. | 0 – 12 |
| **Hidden Objects** |  |  |  |
| Practice round | Practice | Practice round to familiarise participants with the interface and instructions. | – |
| Rounds 1, 2, 3 | Visual processing speed | Participants were required to find a series of objects within a grid populated with distractor objects. | 0 – 9 |
| **Are We There Yet?** |  |  |  |
| Practice round | Practice | Practice round to familiarise participants with the interface and instructions. | – |
| Rounds 1, 2, 3 | Visual learning | In each round, participants were presented with the same route, travelling from one side of a map to the other around various landmarks and were instructed to reproduce the route. | 0 – 36 |
| Round 4 | Visual memory | After a short delay, participants were instructed to reproduce the correct route from memory. | 0 – 12 |
| Round 5 | Recognition memory | Participants were instructed to select which landmarks they remembered seeing along the route presented during the first three rounds. | 0 – 12 |

**Figure 1:**
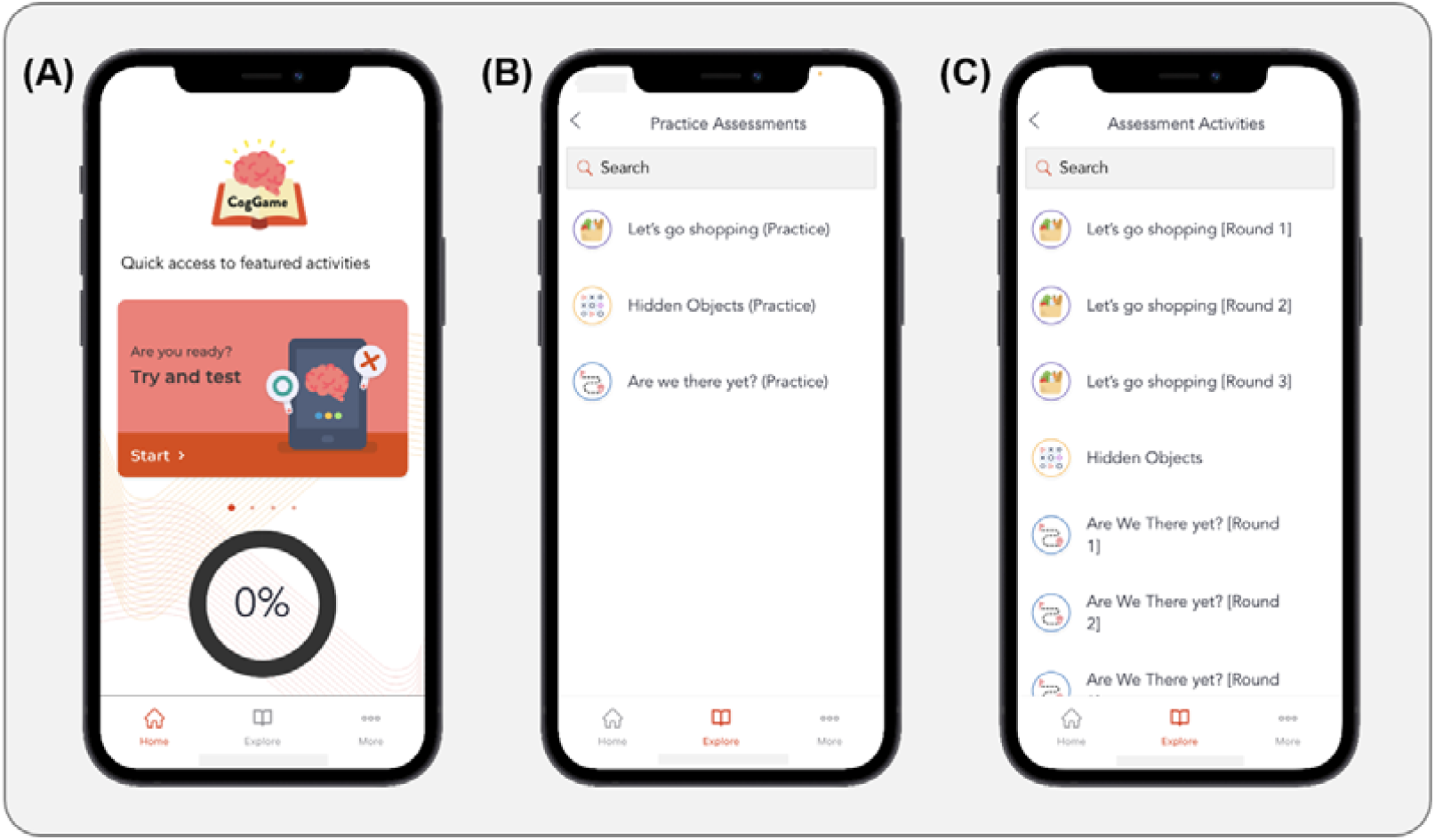
Screenshots of the CogGame interface: (A) Home page. (B) List of practice assessment rounds. (C) List of actual assessment rounds.

**Figure 2:**
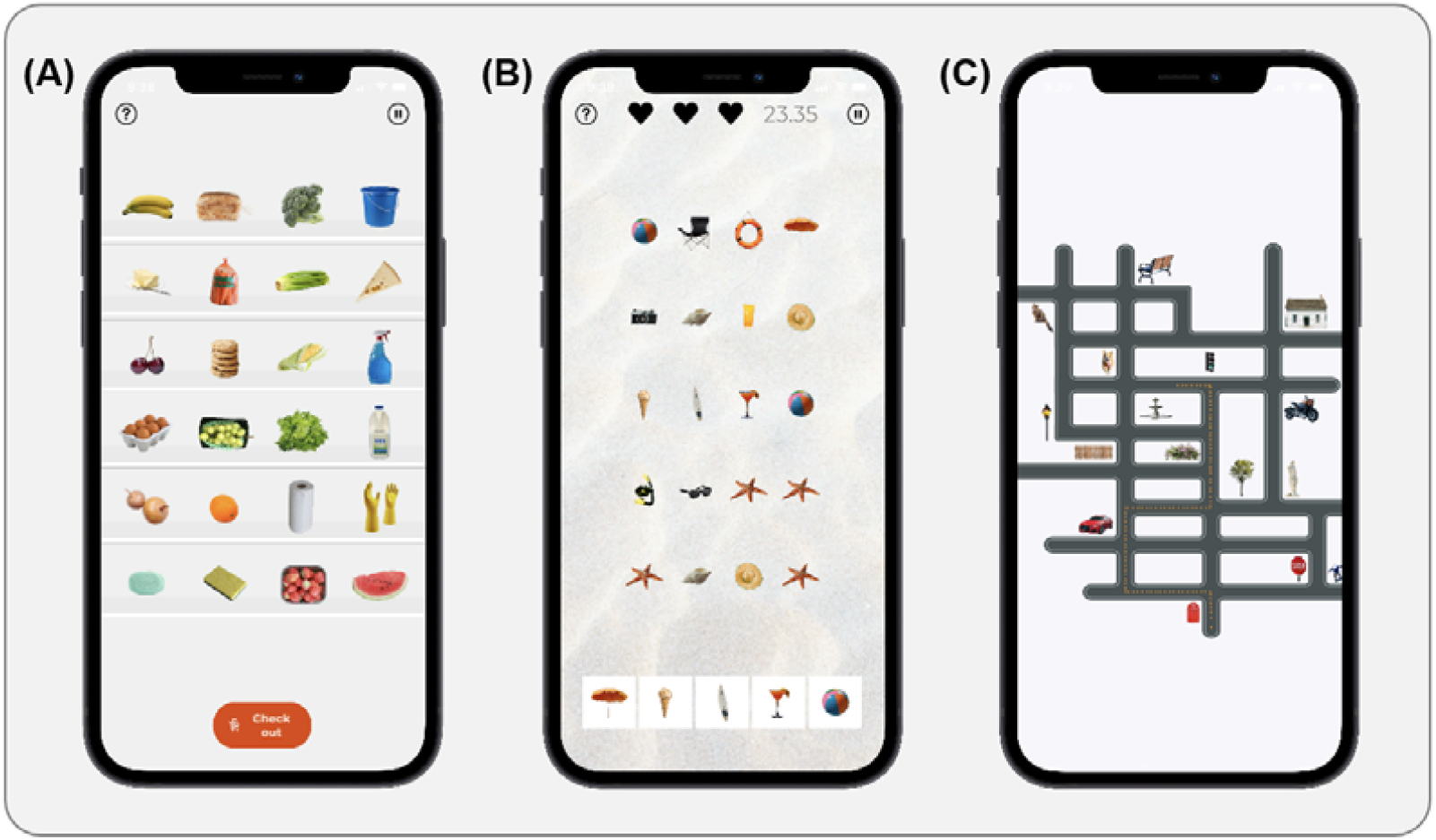
Screenshots of each game in the CogGame app. (A) Let’s Go Shopping. (B) Hidden Objects. (C) Are We There Yet?

#### Sociodemographic variables

Various sociodemographic variables were also obtained via the online questionnaires as well as using linkage data from a previous study the participants had completed, to minimise participatory burden from providing duplicated data with consent. These variables included in the current analysis involve age, gender, highest level of education (secondary school, Certificate Level I-IV, bachelor’s degree, post-graduate degree), location (city or rural), living situation (living with family, significant other, roommate or alone), relationship status (partner or no partner), sexuality (LGBTI or non-LGBTI), physical and mental comorbidities, and whether they were currently taking medication for any mental health conditions.

#### Perceptions of CogGame

Two validated measures were administered during the semi-structured interview to obtain the participants’ perceptions and feedback about CogGame: the System Usability Scale (SUS), and the After Scenario Questionnaire (ASQ). The SUS is a 10-item subjective questionnaire assessing the usability of the app, with each item scored from “1” (strongly disagree) to “5” (strongly agree) (Brooke, 1996). The final calculated score ranges from 0 to 100, with a higher score indicating greater usability and user satisfaction (Brooke, 1996). Cronbach’s alpha of ASQ was 0.78 in the current study. The ASQ is a 3-item subjective questionnaire examining user satisfaction (Lewis, 1991), with each item scored from “1” (strongly agree) to “7” (strongly disagree). The final score is calculated by averaging the three responses, with a lower score indicating greater user satisfaction (range 1 to 7). In addition, open-ended questions (e.g., “Overall, what did you think of the app?”, “How did you find the games?”, “What are your thoughts on the design and layout of the app?”) were utilised during the semi-structured interview to gain deeper insights into the participants’ perceptions of CogGame.

### Statistical Analysis

#### Quantitative Analysis

Data collected via the questionnaires and gamified assessments were coded into numerical values and analysed using IBM SPSS Statistics 26 (SPSS Inc, Chicago, IL, USA). Descriptive statistics were performed to first describe the population characteristics, followed by correlational analyses to determine the cognitive factors associated with suicide risk, as measured by the SIDAS. Significance levels were set at *p*<.05.

#### Thematic Analysis

The qualitative data collected from interviews were analysed using inductive thematic analysis to identify and report the patterns (themes) present in the data (Braun & Clarke, 2006). Specifically, one research team member (CCYS) first coded the interview transcripts using a bottom-up approach and then organised these codes into over-arching themes for each of the key questions asked in the semi-structured interview. A second researcher (JH) then reviewed and refined the codes and themes, with the final themes determined by consensus between the two researchers. These themes were then defined and accompanied by relevant quotes, as outlined in the results in Table 3.

**Table 2:** Descriptive of the participants’ characteristics and their responses to the CogGame

| Variable | Mean | SD |
| --- | --- | --- |
| Age | 21.69 | 2.90 |
| SIDAS score | 9.62 | 9.92 |
| CogGame |  |  |
| Let's Go Shopping |  |  |
| Verbal learning trials<sup>a</sup> | 32.58 | 2.61 |
| Verbal memory round | 11.62 | 0.51 |
| Recognition round | 11.62 | 0.51 |
| Hidden Objects |  |  |
| Visual processing speed rounds | 0 | 0 |
| Are We There Yet? |  |  |
| Visual learning trials | 32.23 | 3.52 |
| Visual memory round | 11.31 | 0.85 |
| Recognition round | 4.08 | 1.50 |
| Variable | N | % |
| Sex |  |  |
| Male | 3 | 23.1% |
| Female | 10 | 76.9% |
| Education level<sup>a</sup> |  |  |
| Diploma, bachelor, or above | 6 | 50.0% |
| Others | 6 | 50.0% |
| Location<sup>a</sup> |  |  |
| City | 8 | 66.7% |
| Rural | 4 | 33.3% |
| Living situation<sup>a</sup> |  |  |
| Living with family or significant other | 9 | 75.0% |
| Others | 3 | 25.0% |
| Relationship status<sup>a</sup> |  |  |
| Partner | 2 | 16.7% |
| No partner | 10 | 83.3% |
| Sexuality<sup>a</sup> |  |  |
| LGBTI | 6 | 50.0% |
| Non-LGBTI | 6 | 50.0% |
| Physical health condition<sup>a</sup> |  |  |
| At least one long-term condition | 4 | 33.3% |
| No long-term condition | 8 | 66.7% |
| Mental health condition<sup>a</sup> |  |  |
| At least one diagnosis | 9 | 75.0% |
| No diagnosed condition | 3 | 25.0% |
| Current mental health medication |  |  |
| Yes | 7 | 53.8% |
| No | 6 | 46.2% |

<sup>a</sup> Sample sizes different due to missing data (n=12).

**Table 3:** Themes emerging from the participants’ responses to the CogGame.

| Theme | Description | n (%) | Examples |
| --- | --- | --- | --- |
| **Engagement** |  |  |  |
| Overall experience | The app was engaging and inviting to use. The games and app features challenged and motivated participants to do well. | 10<br>(76.9) | "It was fun."<br>"I liked that it rewards me as a user, saying "good job" and [with] the badge" |
| Difficulty of the games | The games were not challenging enough. | 8<br>(61.5) | "The games were super easy." |
|  | The games were challenging and satisfying. | 4<br>(30.8) | "It was a good mix of challenges, and the time aspect of the second game adds an extra challenge."<br>"I liked that it was a little bit challenging." |
|  | Some participants did not enjoy being challenged with more difficult games, found them effortful, and were dissatisfied with their performance level. | 4<br>(30.8) | "I found some of them a little stressful."<br>"There was only frustration at not being as good at it as I wanted."<br>"It's just mentally taxing." |
| Target audience | The games may be more appropriate for a different population, such as children or elderly people. | 3<br>(23.1) | "It's got huge potential for testing with children."<br>"I feel like CogGame is something for adolescents or teenagers, or elderly with early signs of cognitive decline." |
| **Functionality** |  |  |  |
| Overall experience | The app was easy to learn to use and navigate. | 13<br>(100.0) | "It's very friendly to the user."<br>"It's easy to get used to and know where to go." |
| Navigation | The transitions between screens could be more efficient. | 9<br>(69.2) | “Going from the practices to the actual assessments could’ve been more logical.” |
| Instructions | Instructions for the games were easy to understand. | 5<br>(38.5) | “The instructions were pretty clear.”<br>“It was very plain, even for non-Science people.” |
|  | Instructions for the games could be improved by adding the goal and purpose of playing the games. | 8<br>(61.5) | “Sometimes I just had to play the games to see how it worked.”<br>“You don’t get feedback about how you went.” |
| ***Aesthetics*** |  |  |  |
| Clean and appealing design | The design of the app interface was clean and appealing, with an aesthetic colour scheme. | 11<br>(84.6) | “I love the interface, the graphic design’s so fun. It’s nice and clean.”<br>“It’s colourful and visually appealing.” |
| Improve design aesthetics | The presentation of the games and the colours or logos used in the app could be more aesthetically pleasing. | 2<br>(15.4) | “Maybe use a more graphical kind of display rather than listing it out.”<br>“It might be worth to play around with different colours, pastels, and logos.” |

## Results

The characteristics of the participants (n=13) and their response to the CogGame app are outlined in Table 2. Majority are female (76.9%), have a degree of diploma, bachelor or above (50.0%), live in city area (66.7%), live with family or significant other (75.0%), have no partner (83.3%), self-identify as Lesbian, Gay, Bisexual, Transgender, Queer and Intersexed (50.0%), have at least one diagnosed mental health condition (75.0%) and no long-term physical health condition (66.7%), and currently take mental health medication (53.8%).

Young adults’ perceptions of the CogGame app were evaluated by the SUS, the ASQ, and the interviews. The mean scores on the SUS and ASQ were 83.65 (SD=9.28) and 1.63 (SD=0.64) respectively, indicating high user-satisfaction and usability. However, one participant responded to question 2 of the ASQ with a score of 6, who preferred a maximum total completion time of 10 to 15 minutes to the actual time of approximately 30 minutes. Apart from this response, all other responses to the three questions of the ASQ were within the range of 1 to 3, revealing an overall satisfaction with the ease of using the app, as well as the instructions and help available when completing the games. Furthermore, thematic analysis of the 13 semi-structured interviews revealed three major themes (engagement, functionality, and aesthetics) regarding the participants’ experience of with CogGame (see Table 3).

Majority of the participants found their experience with the app was engaging and inviting (76.9%). Eight (61.5%) participants thought the cognitive games were easy, while four (30.8%) indicated the games were challenging enough to be satisfying. It is noticeable that four participants (30.8%) suggested that they felt stressful when the difficulty of the games increased. All participants thought the CogGame app was easy to learn to use and navigate in general, although improvement on the transitions between screens (69.2%) and instructions for the games (61.5%) were proposed by some participants. Majority of the participants agreed that the design of the app interface was clean and appealing (84.6%). Only two (15.4%) participants thought the presentation of the games and the colours used in the app could be further improved. Visual learning performance as indicated by the CogGame scores was significantly associated with the levels of the SIDAS scores (see Table 4), with a Spearman’s correlation coefficient value of −0.683 (*p*=.010), indicating poorer visual learning was correlated with more severe suicidal thoughts. No significant relationship between the severity of suicidal thoughts and other cognitive functioning in the current study.

**Table 4:**
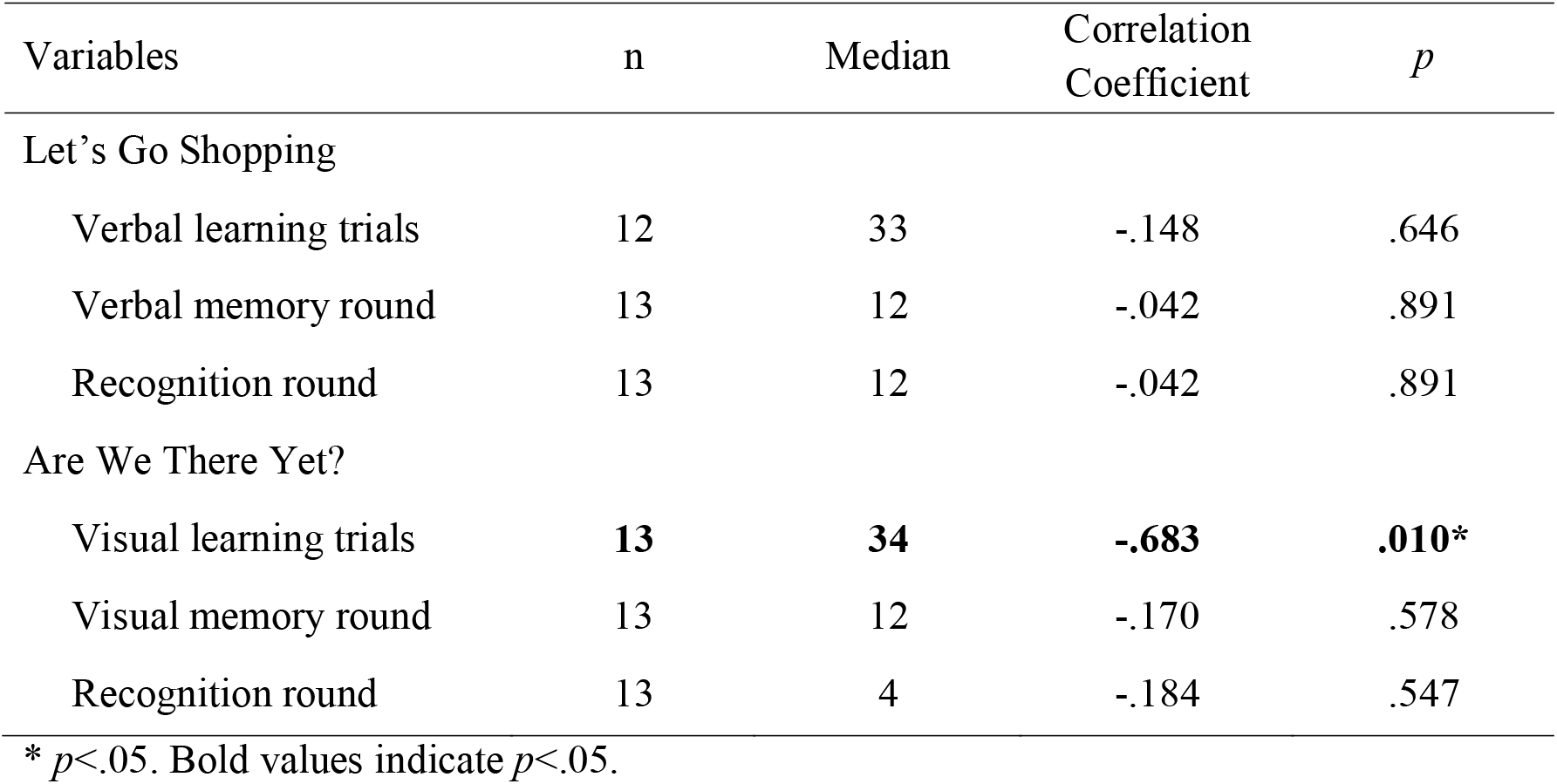
Median and Spearman’s correlation for cognitive functions and SIDAS scores.

| Variables | n | Median | Correlation Coefficient | *p* |
| --- | --- | --- | --- | --- |
| Let's Go Shopping |  |  |  |  |
| Verbal learning trials | 12 | 33 | -.148 | .646 |
| Verbal memory round | 13 | 12 | -.042 | .891 |
| Recognition round | 13 | 12 | -.042 | .891 |
| Are We There Yet? |  |  |  |  |
| Visual learning trials | **13** | **34** | **-.683** | **.010\*** |
| Visual memory round | 13 | 12 | -.170 | .578 |
| Recognition round | 13 | 4 | -.184 | .547 |

\* $p < .05$ . Bold values indicate $p < .05$ .

## Discussion

Overall, positive experiences and high user satisfaction were reported with the use of CogGame, though areas of improvement were noted by the participants, including the need to improve the app’s navigation efficacy and instructions for the games. The exploratory analysis examining the relationship between cognitive functioning and the severity of suicidal thoughts revealed that poorer performance in visual learning was significantly associated with more severe suicidal thoughts in this cohort of young adults.

Thematic analysis of the participants’ semi-structured interviews revealed predominantly positive experiences with CogGame, supported by the scores on the SUS and ASQ scales indicating high usability and user satisfaction with the app. Most participants reported a fun and enjoyable experience and were satisfied with the level of difficulty and challenges offered by the games. Participants also complimented the clean and appealing design of the app, as well as the ease of use and intuitive functionality. These positive experiences demonstrate the high acceptability and promising potential of gamified assessments as a tool in assessing cognitive functioning, with the ability to improve participant engagement and potentially identify high-risk individuals.

Despite the positive feedback, participants also commented on areas for improvement. A common suggestion was to improve the app content by increasing the difficulty of the games to enhance stimulation, as well as including more rounds and variety in the games to maintain participant interest and motivation. A potential way to address this feedback may be through dynamic difficulty adjustment (DDA), a technique to adaptively change a game’s difficulty depending on the player’s performance, thus preventing boredom from games that are too easy or frustration from games that are too hard, consequently maximising the player’s engagement throughout the entire process (Xue, Wu, Kolen, Aghdaie, & Zaman, 2017). Past studies have generally demonstrated greater participant engagement and game experience in commercial (Xue et al., 2017), educational (Sampayo-Vargas, Cope, He, & Byrne, 2013; Shohieb, Doenyas, & Elhady, 2022) and memory training (Araujo, Gonzalez, & Mendez, 2019) games, and thus may also have the potential to enhance motivation with CogGame.

Repairing technical issues, improving the clarity of game instructions, and optimising the efficiency of navigation between game rounds were also noted as possible changes to enhance the usability of the app. Further suggestions included the addition of a progress or outcome report, information about the goal and purpose of the app, as well as a list of helpful resources. Additionally, a few participants also noted that the games seem more appropriate for a different target audience, such as children or the elderly, though this may be because the app was originally designed to target older adults.

The exploratory analyses on the relationship between cognitive functioning deficits measured by the CogGame app and the severity of suicidal thoughts indicate that only the performance on the visual learning task was significantly associated with SIDAS scores. This finding indicates a possible relationship between poorer visual learning performance and severity of suicidal ideation in young adults. Although the lack of significance for the other aspects of cognitive functioning (i.e., verbal learning, visual memory and verbal memory) conflicts with some of the existing findings (Barzilay et al., 2019; Lan et al., 2020; McHugh et al., 2021; Richard-Devantoy et al., 2015), this disparity may due to the relatively small sample size in the current study. A larger-scale study involving young adults with a broader range in the severity of suicidality is recommended to further validate the gamified cognitive assessment and to understand its efficiency in predicting suicidal behaviour.

### Limitations

The current study for the first time presents a promising and engaging alternative to traditional methods of assessing an individual’s cognitive functioning levels in suicidal younger adults. Several limitations need to be addressed regardless the novelty of the study. Firstly, although a small sample size was sufficient to fulfil the exploratory aims of this pilot study, this limited reliable quantitative analyses, thus larger sample sizes are recommended for future studies. Secondly, the findings from the current study were prone to self-selection bias due to the online recruitment method and the voluntary nature of participation, meaning the sample may have consisted of more high-functioning individuals without severe suicidal thoughts or cognitive deficits. Nonetheless, our study provides valuable evidence of numerous advantages of gamified assessments from a participant’s perspective, with data gathered from a unique suicidal young adult population.

## Conclusion

The current study revealed positive experiences regarding the usability, feasibility, and acceptability of CogGame, highlighting the promising potential of gamified assessments as novel, alternative measures of cognitive functioning. Further study is needed to validate these assessments and confirm their accuracy and reliability. This research assisted in developing reliable methods of identifying individuals at suicide risk, thus ultimately improving suicide prevention efforts by directing interventions towards these individuals and their needs.

## Data Availability

All data produced in the present study are available upon reasonable request to the authors

## Acknowledgements

The study was supported by the Bupa Health Foundation Emerging Health Researcher Award to JH. JH is supported by the National Suicide Prevention Post-Doctoral Research Fellowship (RG193218). LM is supported by a joint National Health and Medical Research Council – Australian Research Council Dementia Research Development Fellowship (1109618). The development and build of the CogGame was supported by funding from the University of Sydney-University of New South Wales seed funding scheme: Mental Health and Wellbeing – Early Intervention awarded to HML, JH, VWSC, and LM.

## Data Availability Statement

The data that support the findings of this study are available on request from the corresponding author, JH. The data are not publicly available due to ethics restrictions on the privacy of research participants.

## Conflict of Interest Disclosure

No conflict of interest has been declared by the authors.

## Notes

### Competing Interest Statement

The authors have declared no competing interest.

### Author Declarations

The ethics committee of the University of New South Wales gave ethical approval for this work (HC210432).

